# Integrating Step-down Care in Community-based Paediatric Palliative Services: A Realist Evaluation

**DOI:** 10.1101/2024.03.08.24303949

**Authors:** Zhi Zheng Yeo, Chong Poh Heng

## Abstract

**Background:** Young persons with advanced life-limiting illness living at home have fluctuating and complex needs. Community paediatric palliative care (PPC) is still predominantly specialist-led. This poses a potential care vacuum as medical conditions stabilise and specialised care ceases. One home-based PPC service introduced a step-down care program (COMET) as bridge to continuous yet adaptable support. Above overall effectiveness, how different outcomes are achieved is also investigated for context.

**Methods:** We conducted a realist evaluation, using a mixed-methods case series design to study COMET operations and impact within the community care context (Context->Mechanism->Outcomes). Patient medical records and in-depth interviews with family caregivers and PPC professionals generated rich quantitative and qualitative data for analysis.

**Results:** Of 121 patients under specialist PPC homecare, 18 (14.9%) were enrolled in COMET since November 2020; 12 of these formed individual case studies. Interviews with 15 caregivers and 7 PPC clinicians produced three crucial findings: (i) Ongoing access to specialist care is required for ever evolving complexities; (ii) Continuing support at home is vital for regular management and intermittent emergencies; (iii) COMET harmonizes shifting levels of support within a single unified framework, safeguarding existing rapport and care consistency.

**Conclusion:** Until generalist PPC expertise becomes prevalent outside the hospital setting, novel care models like COMET could plug gaps in community PPC services by offering flexible care options. Continuity of care, efficient resource management, and superior service quality are inherent benefits, if meaningful care tiering through substantive patient assessments are refined further in future iterations.

## Introduction

Individuals with life-limiting and life-threatening illnesses from childhood face numerous challenges, including functional and developmental limitations, recurring symptom episodes, deterioration and truncated lifespans, often not extending beyond early-to-middle adulthood [1]. In many cases, curative treatment is infeasible; hence, these individuals spend much of their lives visiting hospitals for symptom management and life-prolonging interventions, which is associated with substantial trauma and unhappiness [2]. The strain is magnified when the patients are children or adolescents, significantly impacting the well-being of their family caregivers [3].

Consequently, there is a strong preference among these patients and their relatives for receiving palliative care at home or in community settings, where support can extend medical, psychosocial, and spiritual comfort to the entire family unit [4]. Yet, the demand for paediatric palliative care (PPC) in the community outstrips its supply [5], and although its importance is increasingly recognized, typically only those with the most severe illnesses or nearing end of life receive community-based care from specialist PPC providers.

In Singapore’s context, Star PALS by HCA Hospice is the sole specialist-grade community-based PPC service that renders home-based PPC care for children with terminal cancers and non-cancer conditions, and adults who have experienced developmental impairments since childhood due to their illness [6]. Typically, patients continue with the service until death. However, the care pathway becomes less clear for those who reach a temporary state of stability and are perceived to have no outstanding specialist-level PPC needs. Such patients may undergo assessments to determine if they should remain in the service or be discharged [7].

Live discharge from community-based palliative care services is not a well-studied but fairly common phenomenon [8,9]. An audit showed that most adult cases are referred to general practitioners, with very few referred back to hospital; a percentage of cases were eventually re-referred back to the community service [9]. However, this differs from the experience of the PPC service; according to Star PALS discharge statistics, most patients discharged live from the service are referred to hospital care.

Nevertheless, discharge from hospice care has been fraught with challenges and concerns [8]; similarly, departure from the Star PALS PPC tends to go against patient and family preferences, who would prefer to remain under community care. Despite reassurances that re-enrolment into PPC services is possible through hospitals, these patients and families are effectively left to manage care at home independently. For those who reject hospital referrals, they may find themselves without adequate support, and potentially incur significant healthcare-related costs [6,8].

In response to care gaps and client feedback, Star PALS initiated a progressive step-down programme called COMET from November 2020. Modelled after strategies from Intensive Care Units [10,11], the programme targeted those: (i) without pressing specialist PPC requirements; (ii) with longer expected survival; (iii) demonstrating variable care needs; and (iv) lacking appropriate community follow-up options. COMET involved lower-intensity management within specialist PPC services for enrolled patients—reducing consultation (home visits) frequency and access to certain benefits, while preserving crucial crisis assistance, emergency visits, medication access, and remote support. Should a medical emergency arise, rapid re-escalation to full specialist care is available without the need for hospital re-referral. Appendix 1 provides an overview contrasting specialist PPC services with this step-down approach.

The COMET programme’s relatively novel application in community PPC warrants thorough examination regarding its operations, outcomes, and optimal circumstances for effectiveness [12]. A deeper insight into such care models can increase awareness and produce evidence on its potential benefits for patient and family quality of life.

### Objectives

The primary objective of this study is to assess and elucidate the impact of integrating the COMET program into a community-based, specialist-grade PPC service. The overarching research question is: "What are the effects of step-down care on the quality of care for patients with childhood life-limiting illnesses and their families, and under what circumstances, and for whom, does it provide benefits?” The specific aims are as follows:

1. To delineate the characteristics of patients who stand to gain from a step-down care model within paediatric palliative care services.
2. To explore how caregivers and recipients perceive, evaluate, and employ the services offered by COMET according to different needs.
3. To evaluate patient outcomes through participation in the COMET program and contrast these with outcomes observed in those receiving comprehensive specialist PPC services, ultimately to evaluate whether the step-down approach impacts the quality of care delivered.

## Methods

To conduct a thorough analysis of the COMET program, we utilised a realist evaluation approach. This methodology would be suitable at uncovering theories that explain the causative factors behind the changes brought about by a program within its specific contexts [13]. Following this paradigm, we scrutinized (i) the context of the program’s introduction, (ii) the interactions and reactions of stakeholders to the program, and (iii) the resulting outcomes experienced by those involved. Our report adheres to the RAMESES II quality standards for realist evaluations [14].

### Initial Programme Theory

Prior to commencing the evaluation, an initial programme theory was developed based on (i) conversations with key stakeholders – including family caregivers of patients and clinical leads within the PPC service – and (ii) reviews of service usage and pertinent literature on community-based PPC and step-down care model. Our preliminary theoretical framework suggested, “By enhancing continuity of home-based care and guaranteeing access to crucial & urgent support, COMET reinforces home care for PPC patients in Star PALS, thereby alleviating distress and upholding patient well-being.” (Figure 1) The proposed Context-Mechanism-Outcome (CMO) configurations are outlined as follows:

**Figure 1.**
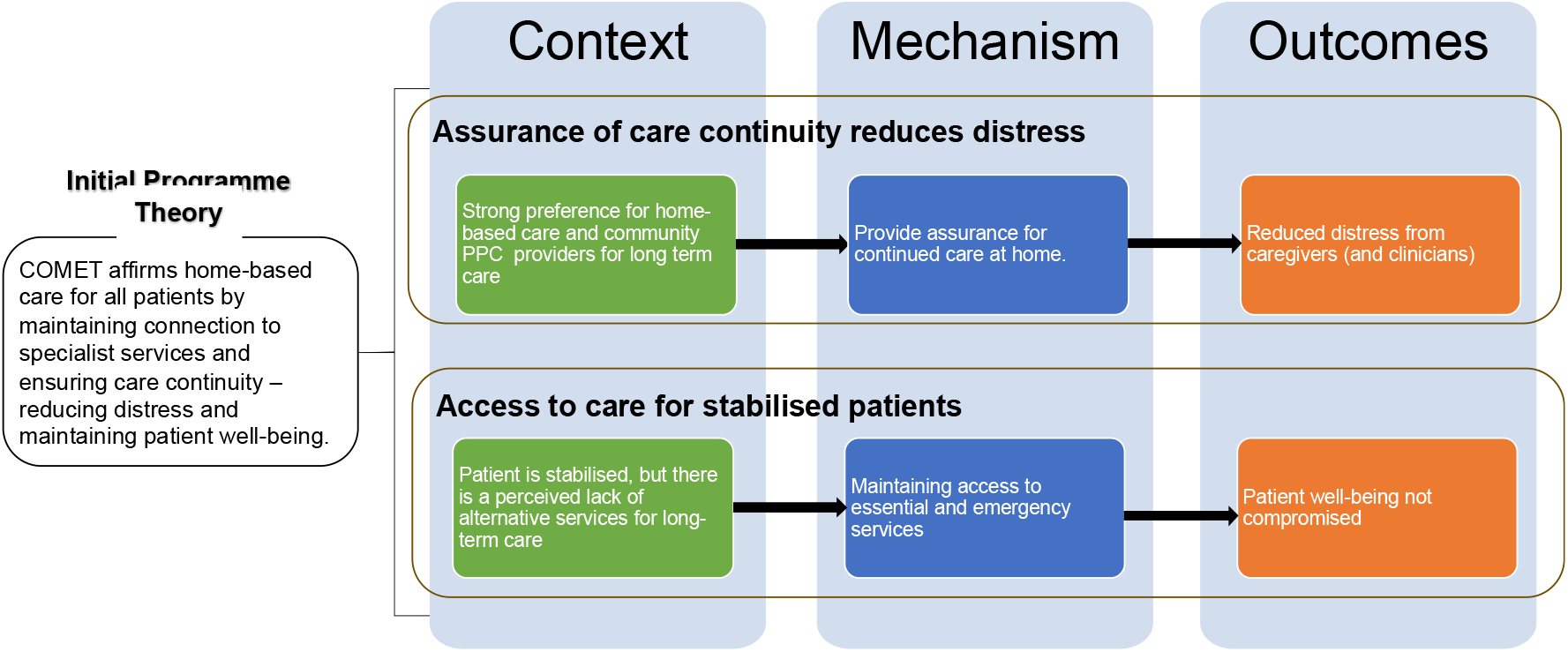
Initial programme theory and hypothesized CMO configurations

#### Care continuity reduces distress

Families prefer home-based care and wish to stay connected with their community PPC team [Context]. COMET intervenes by offering assurance about continuous care at home [Mechanism], which serves to diminish caregiver and clinician distress [Outcome].

#### Access to care for stable patients

There is a scarcity of alternatives for stable, long-term care patients in the community [Context]. Through COMET, patients and families retain access to essential and urgent services within their community settings [Mechanism], such that patient well-being is preserved [Outcome].

### Study design

To delve deeply into the COMET program, we conducted the evaluation using a longitudinal case series, drawing from mixed methods to capture the programme’s complexities and real-world implications.

We targeted the recruitment of 24 patient-cases segmented into three groups: 8 current COMET enrolees, 8 past enrolees who reverted to specialist care, and 8 patients who were discharged from the PPC service alive either before or after COMET’s inception. Each patient was enrolled in the PPC service for at least three months.

We included a wide number of stakeholders of each patient-case —namely, the patient’s family caregivers, primary PPC nurses, social workers, and physicians—for interviews. Stakeholders who met the following criteria were included:

1. Family Caregivers: Chosen if they acted as principal caregivers, lived with the patient, were 21 years or older, and took an active part in daily patient care.
2. Specialist PPC Providers: Valid if they were—or had been—the main nurse, social worker, or physician for any of the participating patients.

A purposive sampling strategy was planned to ensure a variation in contextual elements like patient gender, ethnicity, and socioeconomic status.

### Data Collection & Analysis

Data were collated from patient medical records which provided (i) demographic information, (ii) data on service utilization, and (iii) patient outcomes. The specific types of gathered data are detailed in Table 1.

**Table 1.**
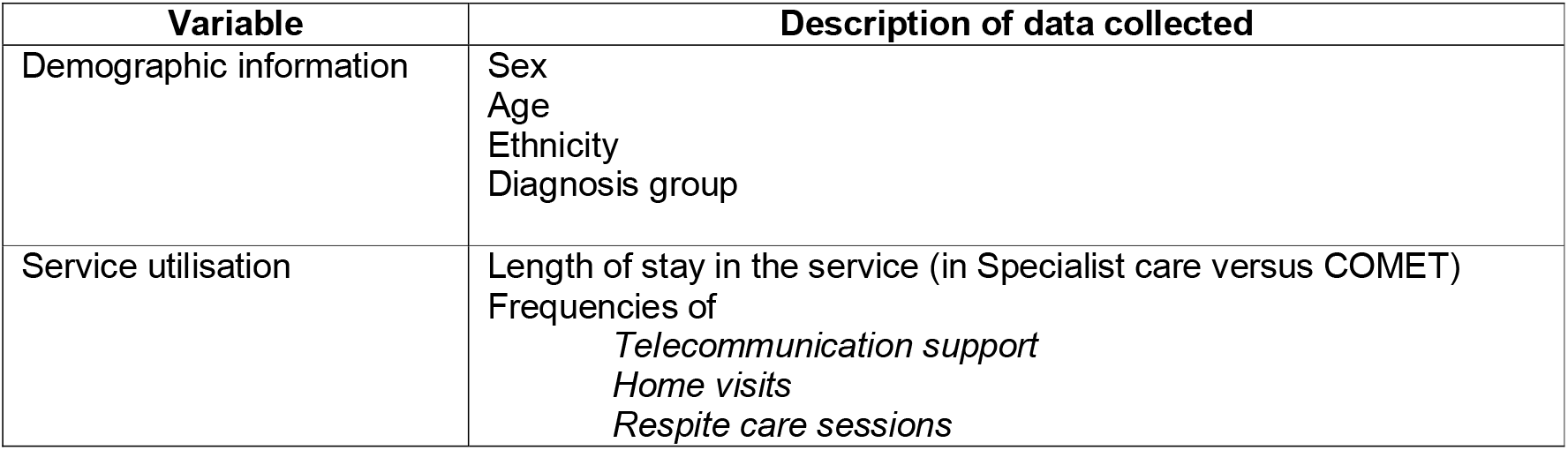
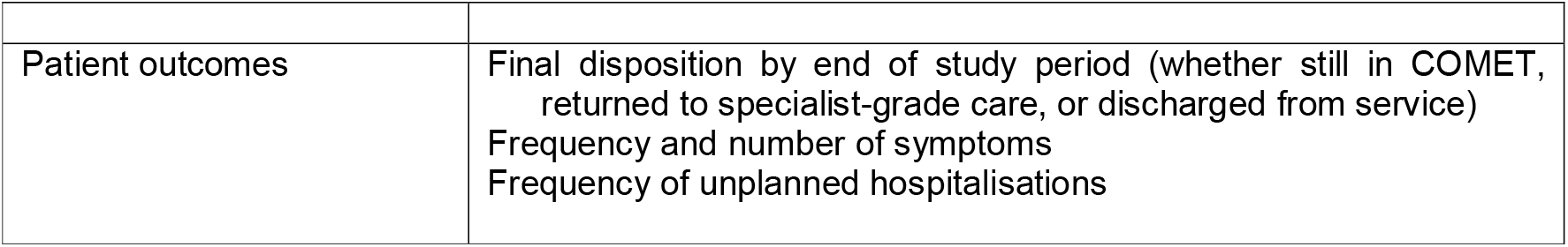
Data collection details.

Quantitative data underwent descriptive analysis to depict demographics and healthcare usage. Additionally, comparative hypothesis testing (α= 0.05) was applied to compare service utilization across two phases for the patients: (i) during the period they received specialist PPC care, and (ii) while they were enrolled in COMET.

Qualitative data was drawn from semi-structured, in-depth interviews with stakeholders. The interviews were conducted by the investigator ZZY via face-to-face or through video conferencing in English, Chinese, or Malay. All interviews were then transcribed in English (translated by ZZY). The transcriptions were subsequently coded using the qualitative data analysis software *QuirkOS*, and analysis was guided by the realist thematic analysis approach outlined by Wiltshire & Ronkainen [15]; multiple inferential modes were leveraged: deductive (using established theory); inductive (driven by data); abductive (seeking the best explanation); and retroductive (pinpointing underlying mechanisms).

Integrating quantitative and qualitative findings involved four principal actions: (i) developing themes and concepts; (ii) linking these with outcomes from both data types; (iii) contrasting results with our initial program theory; and ultimately (iv) finalizing CMO configurations alongside a refined program theory.

Prior to data extraction and conducting interviews, informed consent was obtained from all participants. To preserve confidentiality, pseudonyms (assigned alphabetically based on recruitment dates) replaced patient names, and caregivers’ identities were anonymized during analysis.

## Results

### Case selection

Figure 2 illustrates the selection of cases for the study. Two years between the initiation of COMET and the conclusion of data collection (June 2022), 18 out of a total 121 patients (14.9%) served by Star PALS were enrolled in the program. Of these, 12 consented to participate in the case study, representing just under 10% of the total patient census.

**Figure 2.**
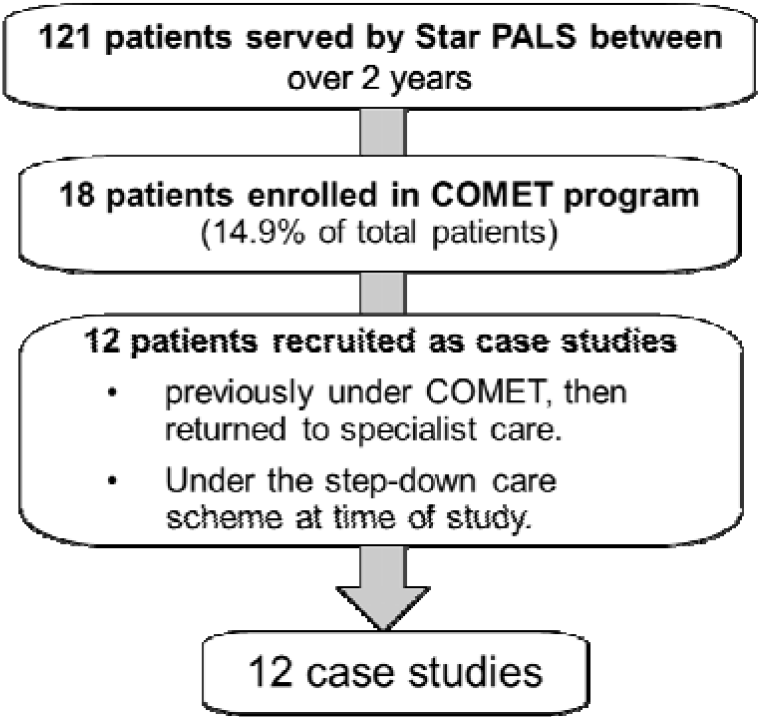
Flowchart of case selection

Patients who had been discharged from Star PALS were approached for recruitment into the study. However, only two family-caregivers (of two discharged patients) consented to participate; others had declined further contact. Interviews with these caregivers provided insights into their experiences with life post-community care, including their transition back to hospital services; however, no direct comparisons were made with patients still in Star PALS or enrolled in COMET. Further details regarding patient cases can be found in Appendix 2.

### Quantitative results

Table 2 displays the demographics and statuses of the 12 patient cases at study end. Most participants were male (75%), with a mean age of 15.9 years (interquartile range: 12–18). All had non-cancer diagnoses and suffered substantial development and functional disabilities. Prior to transitioning to COMET, patients had received specialist PPC services for a mean duration of nearly 30 months, while their enrolment in COMET averaged at about 9.5 months (IQR: 4-17 months). By the study’s close, five cases continued steadily in COMET. One patient was successfully discharged following a liver transplant and no longer needed continuing palliative care services; this individual is an outlier due to no longer meeting palliative care criteria post-operatively. Of the remaining five patients: five re-entered specialist service owing to health crises or deterioration, and one was transferred to a nursing home upon the recognition that home care was no longer tenable.

**Table 2.**
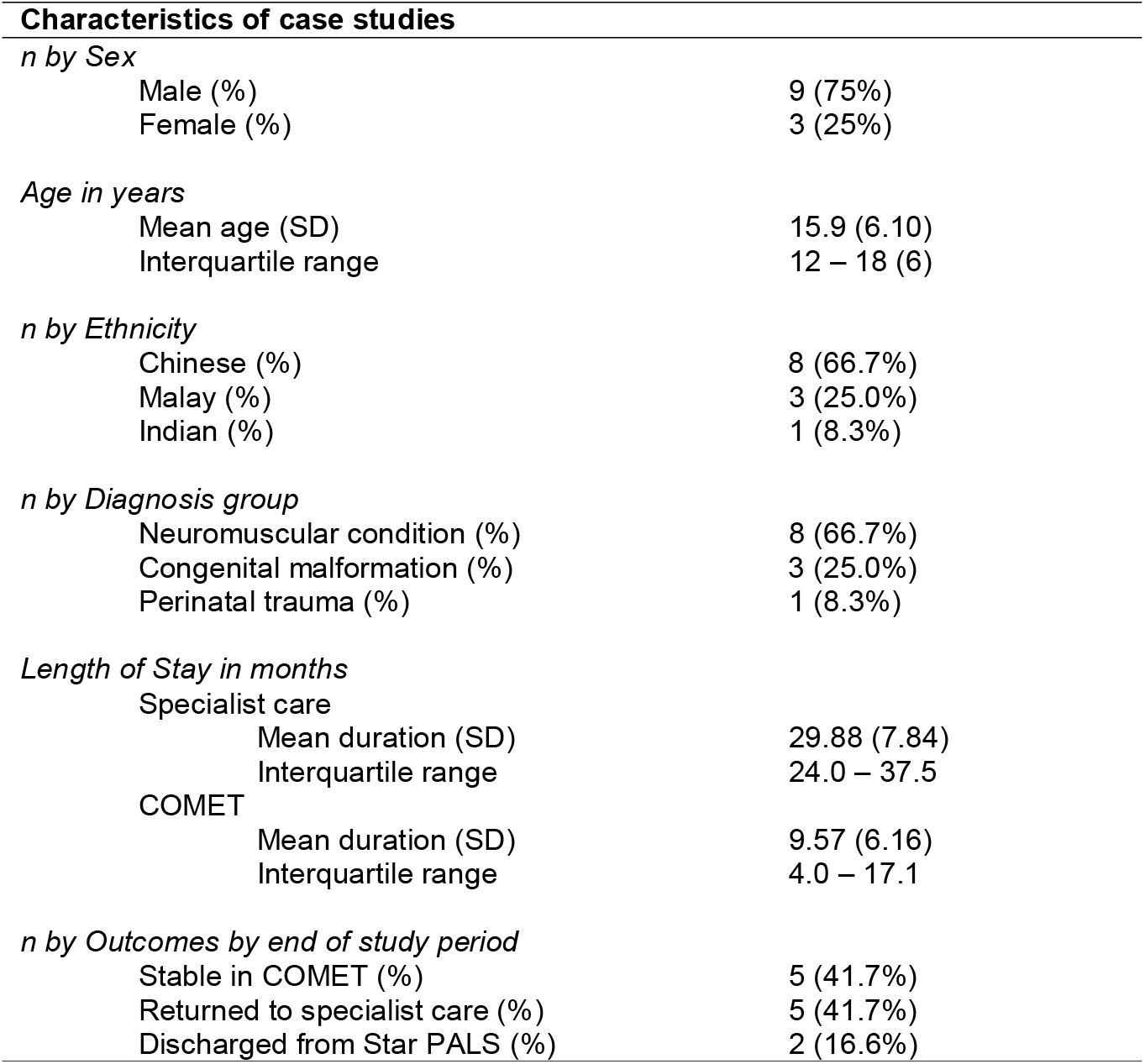
Descriptive characteristics of case studies (N=12)

Table 3 summarizes both service utilization and patient outcome variables from the quantitative analyses. Between both periods, while patients had no difference in monthly frequencies of telecommunications (MD=0.09, p=.864) and respite care sessions (MD=0.11, p=.535), they received 69.1% fewer home visits per month after transitioning to COMET (MD=1.63, p<.001). Overall, the transition from specialist care to COMET was associated with 38% fewer mean contacts (MD=1.82, p=.031). In terms of patient outcomes, Patients reported 85.4% fewer symptoms during their enrolment in COMET, compared to when they were in specialist care (MD=1.19, p<.001). Critically, the transition to COMET was not associated with an increase in the number of unplanned hospitalizations.

**Table 3.** Comparison of cases’ service utilisation and patient outcomes, between the period they were in specialist care versus in COMET (N =12)

| Healthcare utilisation & patient outcomes | Specialist care | COMET | p-value (two-tailed) |
| --- | --- | --- | --- |
| Average contacts per month (SD) | 4.80 (2.01) | 2.98 (1.79) | .031* |
| Telecommunications (SD) | 2.07 (1.44) | 1.98 (1.41) | .864 |
| Home visits (SD) | 2.36 (0.70) | 0.73 (0.35) | <.001* |
| Respite care sessions (SD) | 0.38 (0.69) | 0.27 (0.47) | .535 |
| Average number of symptoms per month (SD) | 1.39 (0.80) | 0.20 (0.20) | <.001* |
| Median Number of unplanned hospitalisations | 2 | 0 |  |
| IQR | 3 (0 – 3) | 1 (0 – 1) |  |
| Maximum | 8 | 1 |  |

\*Test: Paired two-sample t-test; significance at $p < .05$ .

### Qualitative results

Table 4 provides an overview of the demographics of interviewees. In total, interviews encompassed views from 15 caregivers and seven professionals involved in specialist PPC provision—regrettably none of the patients contributed data due to cognitive constraints. Among the caregiver cohort included 8 mothers (including one foster mother) or grandmothers alongside 7 fathers fulfilling principal care responsibilities for these children. Narratives gleaned from three nurses, two medical social workers, and two physicians, provided service and organizational perspectives.

**Table 4.** Demographics of interviewees.

| Characteristics of interviewees | Caregivers (n =15)* | Providers (n =7) |
| --- | --- | --- |
| *n by Sex* |  |  |
| Male (%) | 7 (46.6%) | 2 (28.6%) |
| Female (%) | 8 (53.3%) | 5 (71.4%) |
| *n by Ethnicity* |  |  |
| Chinese (%) | 11 (66.7%) | 7 (100%) |
| Malay (%) | 3 (25.0%) | - |
| Indian (%) | 1 (8.3%) | - |
| *n by Roles* |  |  |
| Mother (including 1 foster mother) | 7 (46.6%) | - |
| Father | 6 (40.0%) | - |
| Grandmother | 2 (13.3%) | - |
| Nurse | - | 3 (42.9%) |
| Physician | - | 2 (28.6%) |
| Medical Social Worker | - | 2 (28.6%) |

% calculated as proportion of group total (caregivers or providers)
\*Two interviewees were caregivers of patients who were discharged from the service before the COMET program was introduced.

From the qualitative data, key themes were extracted to highlight the experiences of stakeholders regarding caring for the child within the community, as well as their experiences with COMET:

### The community care landscape for PPC

#### Inevitable deterioration & increasing care needs

The experience of children with life-limiting illness is characterised by a clear downtrend in health, and growing needs to address emerging symptoms and care needs:

> *“…Compared to [previous years], all her-what you call it-went down [to] all the muscles, getting weaker and weaker. Now she can only move just few-few fingers and just toes only. Yeah, other than that, she won’t move*.*”*
>
> *“…the degeneration is pretty-it’s very gradual. You don’t really see it on consistent day-to-day basis, but… I remember recalling that initially- you know, you really grasp at straws that you want to do what you can, right?”*

#### Limited and fragmented care options

While ideally there would be an ecosystem of healthcare services that can meet their needs, all stakeholders commented that the existing system was lacking in its ability to serve “medically complex” children in the community:

> *“…in Singapore, unlike some of the countries in the West, we do not have a very rigorous kind of home support services; medical-interprofessional home support services for children with medically complex needs. So, that is very challenging*.*”*

#### Unresponsive healthcare

Moreover, there was a perception that the healthcare system did not fully comprehend the needs of the families, which led to gaps and delays in care provision. Caregivers recounted how community healthcare providers would refer their child to hospital, even for minor ailments. Even in hospitals, caregivers described difficult experiences:

> *“Before Star PALS, whenever he was sick, I would just go to A&E [Accidents & Emergencies]. Because when I went to the [general practitioner], they did not want. Private doctors, they don’t want-they told me to go to hospital. If I went to the polyclinic, they would give me a letter, and send me to hospital… I think it’s because they saw that he has many problems, so it would be difficult to treat him. Therefore, they give a letter, and tell me to go to hospital*.*”*

> *“…then [when] they reach the hospital, sometimes there might be a delay. So not only compromised patients’ medical condition, the health condition but also is very stressful for the family”*

#### Reluctance for discharge

Hence, though a child may appear medically stable, families and providers may be reluctant to discharge the child from the specialist service, as it exacerbated these gaps:

> *“…But suddenly [after discharge], I felt that I was-I may not be able to-I can take care of the functional things, but then medically, if things crop up, I don’t have anybody to talk to*.*”*

> *“Let’s say, Star PALS is discharging this patient or moving the patient to a ‘very stable’ group, then whether do they have the resources they need if, let’s say things happen… Yeah, so if let’s say it’s a condition that is very uncertain, I can see why people are more reactive to discharging the patient*.*”*

#### Specialist-role versus community care

Given these logistical, interpersonal, and contextual factors, healthcare providers face a dilemma between the pragmatic need to maintain their specialist role, versus fulfilling an ethical desire to serve more children with life-limiting illness living within the community:

> *“If we keep them, whether we are able to serve the other people who need us more during that period time? Yeah, so this is a struggle*.*”*

> *“While Star PALS tries to meet these areas or gaps in the healthcare system of support for our families-that it does not detract from our main service, expertise, and specialty. Because we are special specialist palliative care service*.*”*

### Implementation of COMET

#### Care continuation for softer transition

COMET was described as a model of care to achieve care continuation and afford patients a softer transition to less intensive care. The sentiment was that it achieved the dual objectives of ensuring patients’ wellbeing, while being a more sustainable mode of service:

> *“This step-down can be rendered by the organic team and therefore, also prevents the people [families] from feeling abandoned- and also that sometimes very problematic transition between services. And of course, potentially softens the impact of being discharged from the service… Done in good faith with the right intention-both for- you know, service sustainability as well as accountability, which is safe-you know, safe as in preventing people from falling between the cracks,”*

#### Adjusting to the child’s needs

For healthcare providers, COMET was experienced as a system that allowed for more nuanced evaluations and decision-making, to better meet the needs and preferences of the child and their family caregivers:

> *“When you reduce a problem to just small, medium, big, and then cut a certain-set a certain threshold, beyond which-okay, I will pick up the case, or below which I will not. I find that problematic when dealing with the vagaries of symptoms at different timepoints in the child’s illness trajectory… But to recognize that they are caveats that-actually, we need to consider, and perhaps leave some room for a decision on a case-by-case basis- that’s when potentially some of the more nuanced-more granular, detailed evaluation can happen*.

#### Consistent care enhances confidence

For caregivers, the highlight of COMET is the perception that important medical services will be maintained and consistent; this lends them greater confidence in their ability to keep the child at home:

> *“We still feel that for her kind of condition, it’s always good to have to constantly have that kind of support, because for her things are just very unpredictable and a lot of times when things suddenly happen it can be quite serious*.*”*

#### Flexible approach to care

After an initial period of adjustment, the PPC providers reflected the flexibility to rapidly adjust the patient’s care (between COMET and CORE) based on prevailing needs was an important contributor to an overall sense of responsiveness. This also helped with the perception that keeping additional patients was not a significantly greater workload:

> *“I mean in terms of workload, of course it has lightened-It has lightened certain level of workload because for-for some patients that I really feel that don’t really be visit frequently, and they’re parked in COMET, I think it helps a lot like knowing that we don’t have to really abide by the once-a-month kind of visit*.*”*

> *“And so far, for the few patients we transferred them back to the core (service), they were also quite appreciative of that, although we transfer them to core or to COMET group, the care was never being stopped*.*”*

### Impact of COMET

#### No compromise to care

Despite the lowered intensity of care rendered, caregivers did not perceive any compromise to their quality of life:

> *“I think the weekly visit is not always necessary. Because our children aren’t always sick, they can be well*.*”*

> *“I don’t mind [being in COMET], you can come once a month or once in two months, but not once in three months. I prefer to see them on a regular basis, but once in three months to me is quite wide*.*”*

However, a few caregivers did highlight that their child experienced periodic episodes of deterioration, and that responsiveness to their situations remains paramount:

> *“…when they talk about discharging her [or putting her in COMET], something will happen. Something happened and is something new to us, to the team. So we have to manage her again, like when they first talk about-example, like you know, going to discharge her because she was stable, then ended up she needed a trache[ostomy]…”*

#### Maintaining a lifeline

A major component of COMET is the maintenance of access to basic and emergency support for patients placed under COMET. To stakeholders, this was perceived as creating a lifeline for specialist support when needed:

> *“I’m still throwing them into sea in some sense, but with a very long line hooking onto their ship. So-it provides reassurance to me as a nurse, that’s one thing. And then I could see that it provides reassurance to the family as well*.*”*

Caregivers echoed the sentiment, highlighting that access to rapid advice and updates was often most relevant to their needs:

> *“As the child’s condition stabilizes, the frequency of nurse visits can decrease- and let them visit those with higher needs more. But I think that it is good if we can still contact the nurses through WhatsApp… once every 3 months is okay*…*”*

### Synthesis: CMO configurations

The COMET program has been met with approval by stakeholders due to its perceived enhancement of home-based care. By synthesizing quantitative and qualitative data, the initial program theory was redefined, highlighting additional subtleties and contexts. This led to the development of the following Context-Mechanism-Outcome (CMO) configurations (Figure 3):

**Figure 3.**
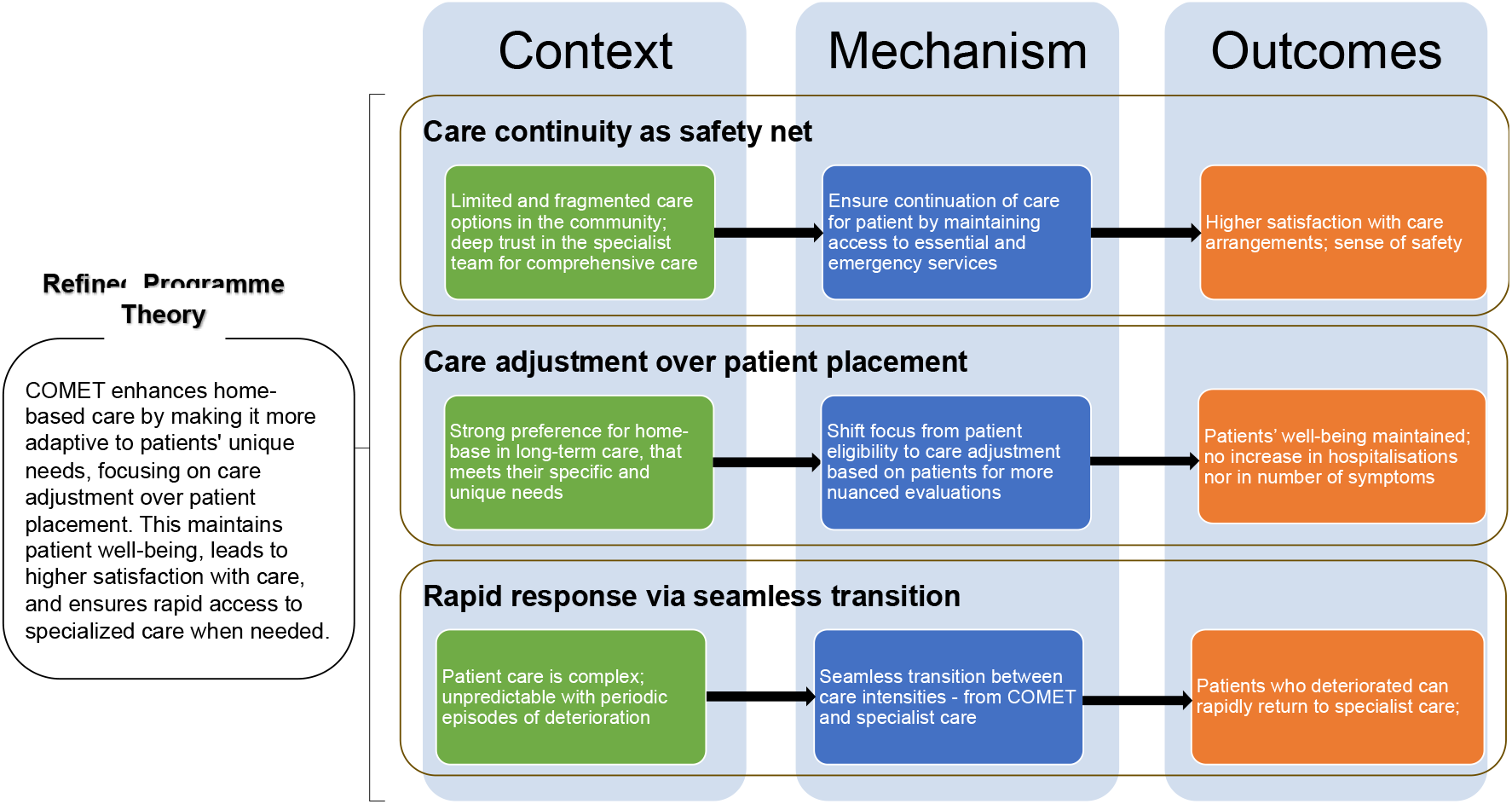
Refined programme theory

#### Care Continuity as Safety Net

In an environment often limited by fragmented healthcare options and a perceived insensitivity of non-specialist services to their needs, individuals with life-limiting illnesses from childhood face significant care gaps and delays [Context]. COMET is viewed as preserving access to critical and emergency services. Families and caregivers draw confidence from the assurance that comprehensive home care will continue [Mechanism], resulting in diminished distress [Outcome].

#### Care Adjustment over Patient Placement

Stakeholders reaffirmed a clear preference for tailored home-based care that addresses patients’ individual and specific needs over a prolonged period [Context]. The focus shifts from the patient’s care setting to customizing the care they receive. Such nuanced care planning strengthens bonds between healthcare providers and families [Mechanism], leading to increased levels of satisfaction with the care arrangement [Outcome].

#### Seamless Transitions between Care Levels

The program further revealed intricacies associated with managing the complex and fluctuating conditions of this patient group, plagued by intermittent episodes of deterioration and uncertainty [Context]. COMET facilitates smooth and rapid transitions between specialist and step-down care as needed, which stakeholders recognized as a responsive system adapting swiftly to changes in patient health [Mechanism]. This flexibility appeared to support patient well-being without noticeably sacrificing quality of care [Outcome].

## Discussion

The COMET program effectively met the needs of a subset of paediatric palliative care patients who can be described as “medically fragile.” These individuals, ultimately susceptible to health fluctuations despite stabilization, benefit from continued at-home care from specialists at a reduced intensity, alleviating caregiver anxiety by ensuring access to essential services. The recurrence of deterioration in many patients underscores the essential nature of such retention in the service. Half of COMET’s patients reverted to intensive specialist care, driven by declining health status or urgent medical needs—underscoring an ongoing necessity for vigilant selection and monitoring processes regarding participant inclusion within such programs.

Home-based care serves as a vital element of consistent, quality healthcare delivery. Palliative care provided in the home is acknowledged for reducing healthcare usage and costs, benefits widely recognized by all stakeholders. Nevertheless, the challenge lies in fulfilling this need without compromising equity; this encompasses decisions on whether to discharge stable but vulnerable patients or prioritize incoming patients requiring more immediate specialist attention. Notably, discharged patients are not cured outright; they’re often situated in a period of uncertainty— highlighted by caregivers who must revert to hospital resources for assistance that was once managed by community services.

The study provides insights into the challenges of long-term care for children with chronic illnesses with no prospects of recovery or cure. The COMET program emerges as a possible solution to this persistent concern, building on existing hospital-at-home models [11] and emphasizing tailored service provisions over location. As a unique iteration within community-based palliative care, COMET supports patients’ needs *in situ*, notwithstanding their current health state. Ultimately, despite Star PALS’s specialist service label, families perceive the service as integral and holistic – a service “with them until the end.” In the meantime, community-based specialist PPC services may need to revaluate their roles within broader patient care landscapes [4], possibly expanding their remit despite their specialist identity until alternative services can offer comparable generalist support.

The COMET model’s prime advantage is its flexibility to modulate care intensity and balance resources effectively. Paediatric palliative needs are dynamic and uncertain [6]; hence a system facilitating varying levels of care intensity may be more suitable than rigid categorizations delineating ’sick/unstable’ from ’stable/well.’ This is notably pertinent for non-cancer conditions characterized by less predictable trajectories with periods of relative stability that challenge the appropriateness of specialist palliative care, contrasting traditional practices which cater mainly to cancer-related trajectories [18].

From this study’s insights, several recommendations have emerged:

1. A shift away from conventionally rigid ’sick/unstable’ versus ’stable’ patient classifications within community care, as this does not always translate into effective or fair service delivery [1].
2. Embrace a care model focusing on adjusting care intensities rather than moving patients between different settings [11], fostering continuity in patient-provider relationships and enabling responsive caregiving with fewer administrative concerns.
3. Implement comprehensive assessment processes coupled with deep engagement with patients and families to support equitable care scaling and minimize perceptions of diminished quality of life and care. Standardized palliative assessment tools can guide the tailored scaling of provided services [7].

### Strengths & limitations

The findings of this realist evaluation should be considered within the context of the strengths and limitations of the study. One strength is the timing of evaluation—in the early stages of the program’s development and implementation, which allows for close monitoring and feedback for ongoing refinement. However, recruitment proved challenging, particularly with patients who had been discharged from the service as their families were reluctant to maintain contact. Consequently, the intended sample size of 24 patients was not met. Nonetheless, this limitation was offset by a comprehensive longitudinal approach that captured multiple data sources, including healthcare utilization figures, clinical outcomes, and qualitative feedback from stakeholders. Future studies might aim to secure a larger cohort for analysis and consider conducting comparative outcome research on those who remain under COMET versus patients discharged from community-based palliative services.

Another core aspect of the study is the diverse set of case studies that include patients from varying socio-economic statuses, allowing for rich data collection and potential generalizability across different patient demographics. Despite this intent for diversity, currently only non-cancer patients are enrolled in COMET. Future initiatives could look to expanding enrolment to include stabilized cancer patients as part of step-down care.

Future research could extend the scope of investigation by applying more experimental designs to directly contrast the efficiency and impact of this step-down care model with alternative approaches. Broadening the evidence base to include perspectives from other essential stakeholders—like policymakers, institutional collaborators, and additional community-based providers—would also augment insights into healthcare delivery for children with severe and life-limiting conditions. This multi-faceted input plays a pivotal role in shaping well-rounded healthcare systems that account for diverse needs and scenarios common in paediatric palliative care.

## Conclusion

In conclusion, COMET expands the scope of specialist paediatric palliative services through the incorporation of step-down care, effectively addressing a critical gap in the community care framework. Our study reaffirms that the heterogeneous healthcare needs of paediatric palliative patients cannot be met with a rigid approach. This reinforces the need for continuous development of community capabilities that allow children to remain in the comfort of their homes, and to mitigate the abrupt cessation of services upon discharge—a reality recognized by stakeholders.

(Word Count: 4,540; 3,580 without quotes)

## Data Availability

All data produced in the present study are available upon reasonable request to the authors

## Declarations

### Ethics approval

This study was reviewed by the SingHealth Centralised Institutional Review Board (CIRB) on 30 August 2021 and determined not to require further ethical deliberation as a program evaluation study (Reference number 2021/2560).

### Contributions of the authors

All authors contributed to the drafting, review, and completion of the manuscript.

## Acknowledgements

The author would also like to acknowledge the families who agreed to participate and share their experiences, as well as the team from Star PALS for their inputs and participation, without which this evaluation would have been impossible.

## Data availability statement

The data that support the findings of this study are available upon reasonable request. Interested researchers may access deidentified qualitative data and aggregated statistical information from HCA Hospice. Requests for data should be directed to the research team via email at. Permission for data reuse can be granted following a discussion and agreement with the authors of the study.

